# Trends in non-cigarette tobacco smoking in England: a population survey 2013-2023

**DOI:** 10.1101/2023.11.24.23298982

**Authors:** Sarah E. Jackson, Lion Shahab, Jamie Brown

## Abstract

**Background:** The UK Government intends to implement a ‘smokefree generation’ policy prohibiting the sale of all tobacco products to people born after 2008. National surveys provide comprehensive data on cigarette smoking, but little is known about patterns of non-cigarette tobacco smoking across key population groups.

**Methods:** Using data from a nationally-representative cross-sectional survey of adults (≥18y) in England, collected monthly between September-2013 and September-2023 (*n*=196,721), we estimated time trends in non-cigarette tobacco smoking prevalence, overall and by age, gender, occupational social grade, region, ethnicity, and vaping status. Interviews were conducted face-to-face until March-2020 and via telephone thereafter.

**Results:** From September-2013 to September-2023, there was a non-linear increase in non-cigarette tobacco smoking prevalence (from 0.36% to 1.68%; PR=4.72 [95%CI=3.43-6.48]). Prevalence was relatively stable up to February-2020 (at an average of 0.46%), then increased sharply at the start of the Covid-19 pandemic, to 0.90% [0.82-0.99%] in March-2020. This was followed by a steadier rise, peaking at 1.97% in May-2022, before falling slightly to 1.68% by September-2023. As a result, in 2022/23, one in ten smokers (10.8% [9.64-12.0%]) used non-cigarette tobacco. This rise was observed across all subgroups but was most pronounced among younger adults (e.g., reaching 3.21% of 18-year-olds vs. 1.09% of 65-year-olds). Prevalence was consistently higher among men (2.17% in September-2023 vs. 1.07% women) and current vapers (4.71% vs. 1.25% non-vapers).

**Conclusions:** While exclusive use of non-cigarette combustible tobacco remains rare among adults in England, it increased at the start of the Covid-19 pandemic (at the same time as survey methods changed) but subsequently continued increasing steadily until May-2022. As of September-2023, there were ∼772,800 adult non-cigarette tobacco smokers in England; around five times more than a decade earlier. The rise in prevalence differed by age, with a more pronounced rise leading to higher prevalence among younger than older ages.

**What is already known on this topic:** There is good evidence from nationally-representative population surveys on the prevalence and patterns of cigarette smoking in England. Less is known about use of other combustible tobacco products.

**What this study adds:** Prevalence of non-cigarette tobacco smoking has risen substantially since the start of the Covid-19 pandemic, particularly among younger adults. As a result, one in 10 smokers in England now does not smoke cigarettes at all but smokes some other form of combustible tobacco.

**How this study might affect research, practice or policy:** The UK Government is planning to ban the sale of tobacco products to those born after 2008. The inclusion of non-cigarette combustible tobacco products under this policy is likely to be important for achieving the greatest reduction in youth uptake of tobacco smoking, as it would ensure young people who are unable to legally buy cigarettes do not buy other combustible tobacco products that are similarly harmful to health.

## Introduction

In October 2023, the UK Government announced its intention to implement a ‘smokefree generation’ policy in England, which would progressively increase the age of sale of tobacco products such that anyone born on or after 1 January 2009 would never legally be able to buy tobacco.^1^ Non-cigarette tobacco products, such as pipes, cigars, and shisha, would be included under this proposed policy.^1^

There is good evidence from representative population surveys on the prevalence and patterns of cigarette smoking in England,^2^ providing a clear picture of who would be targeted by this policy. Such surveys suggest the exclusive use of other non-cigarette combustible products was relatively rare in England through to 2015,^3^ but little is known about the latest numbers who would be affected by the new policy. It is particularly relevant to assess the latest data because prevalence may have changed substantially in the context of a rapidly evolving nicotine market, the Covid-19 pandemic, and other major socioeconomic shifts in the last five years.

Understanding how prevalent non-cigarette tobacco smoking is, and how this is changing over time, is important for monitoring purposes and for informing and evaluating policy. The Smoking Toolkit Study (a representative, cross-sectional survey) collects data on non-cigarette tobacco smoking among adults in England each month. This study aimed to use data collected over the past decade to address the following research questions:

1. What is the overall prevalence of non-cigarette tobacco smoking among adults in England, and how does this vary by sociodemographic characteristics and vaping status?
2. How has the prevalence of non-cigarette tobacco smoking changed between 2013 and 2023?
3. Have trends over this period varied by sociodemographic characteristics and vaping status?

## Method

### Pre-registration

The study protocol and analysis plan were pre-registered on Open Science Framework (https://osf.io/w7f3m/). In addition to our planned analyses, we calculated the absolute and relative change in the prevalence of non-cigarette tobacco smoking across the whole time series and estimated the number of adult non-cigarette tobacco smokers in England in September 2013 and September 2023 (see *Statistical analysis* section for details). We also added an unplanned segmented regression analysis to examine whether the start of the Covid-19 pandemic (and change in the mode of data collection, from face-to-face to telephone interviews) was associated with a step-level change in non-cigarette tobacco smoking and a change in trend.

### Design

Data were drawn from the ongoing Smoking Toolkit Study, a monthly cross-sectional survey of a representative sample of adults aged ≥16 years in England.^4^ The study uses a hybrid of random probability and simple quota sampling to select a new sample of approximately 1,700 adults each month. Comparisons with sales data and other national surveys indicate that key variables including sociodemographic characteristics, smoking prevalence, and cigarette consumption are nationally representative.^4,5^

We used data collected from September 2013 through September 2023 (the most recent data at the time of analysis). Data were initially collected through face-to-face computer-assisted interviews. However, social distancing restrictions under the Covid-19 pandemic meant no data were collected in March 2020 and data from April 2020 onwards were collected via telephone. The telephone-based data collection used broadly the same combination of random location and quota sampling, and weighting approach as the face-to-face interviews and comparisons of the two data collection modalities indicate good comparability.^6–8^

### Population

We used data from adults aged ≥18 years. Because data were not collected from 16- and 17-year-olds between April 2020 and December 2021, we restricted our sample to those aged ≥18 for consistency across the time series.

### Measures

#### Smoking status

was assessed by asking participants which of the following best applies to them:

a. I smoke cigarettes (including hand-rolled) every day
b. I smoke cigarettes (including hand-rolled), but not every day
c. I do not smoke cigarettes at all, but I do smoke tobacco of some kind (e.g., pipe, cigar or shisha)
d. I have stopped smoking completely in the last year
e. I stopped smoking completely more than a year ago
f. I have never been a smoker (i.e., smoked for a year or more)

Those who responded *a-c* were considered current smokers. Those who responded *c* were considered non-cigarette tobacco smokers and those who responded *a* or *b* cigarette smokers.

#### Age

was modelled as a continuous variable using restricted cubic splines (see statistical analysis section). We also provided descriptive data by age group (18-24/25-34/35-44/45-54/55-64/≥65).

#### Gender

was self-reported as man or woman. In more recent waves, participants have also had the option to describe their gender in another way; those who identified in another way were excluded from analyses by gender due to low numbers.

#### Occupational social grade

was categorised as ABC1 (includes managerial, professional, and upper supervisory occupations) and C2DE (includes manual routine, semi-routine, lower supervisory, and long-term unemployed).

#### Region in England

was categorised as North, Midlands, and South.

#### Ethnicity

was categorised as white and minority ethnic group. Data on ethnicity were not collected between April and August 2020.

#### Vaping status

was assessed within several questions asking about use of a range of nicotine products. Current smokers were asked ‘Do you regularly use any of the following in situations when you are not allowed to smoke?’; current smokers and those who have quit in the past year were asked ‘Can I check, are you using any of the following either to help you stop smoking, to help you cut down or for any other reason at all?’; and non-smokers were asked ‘Can I check, are you using any of the following?’. Those who reported using an e-cigarette in response to any of these questions were considered current vapers.

#### Statistical analysis

Analyses were done in R version 4.2.1. The Smoking Toolkit Study uses raking to weight the sample to match the population in England. This profile is determined each month by combining data from the UK Census, the Office for National Statistics mid-year estimates, and the annual National Readership Survey.^4^ The following analyses used weighted data. We excluded participants with missing data on smoking status. Missing cases on other variables (see **Table 1** for details) were excluded on a per-analysis basis.

**Table 1.**
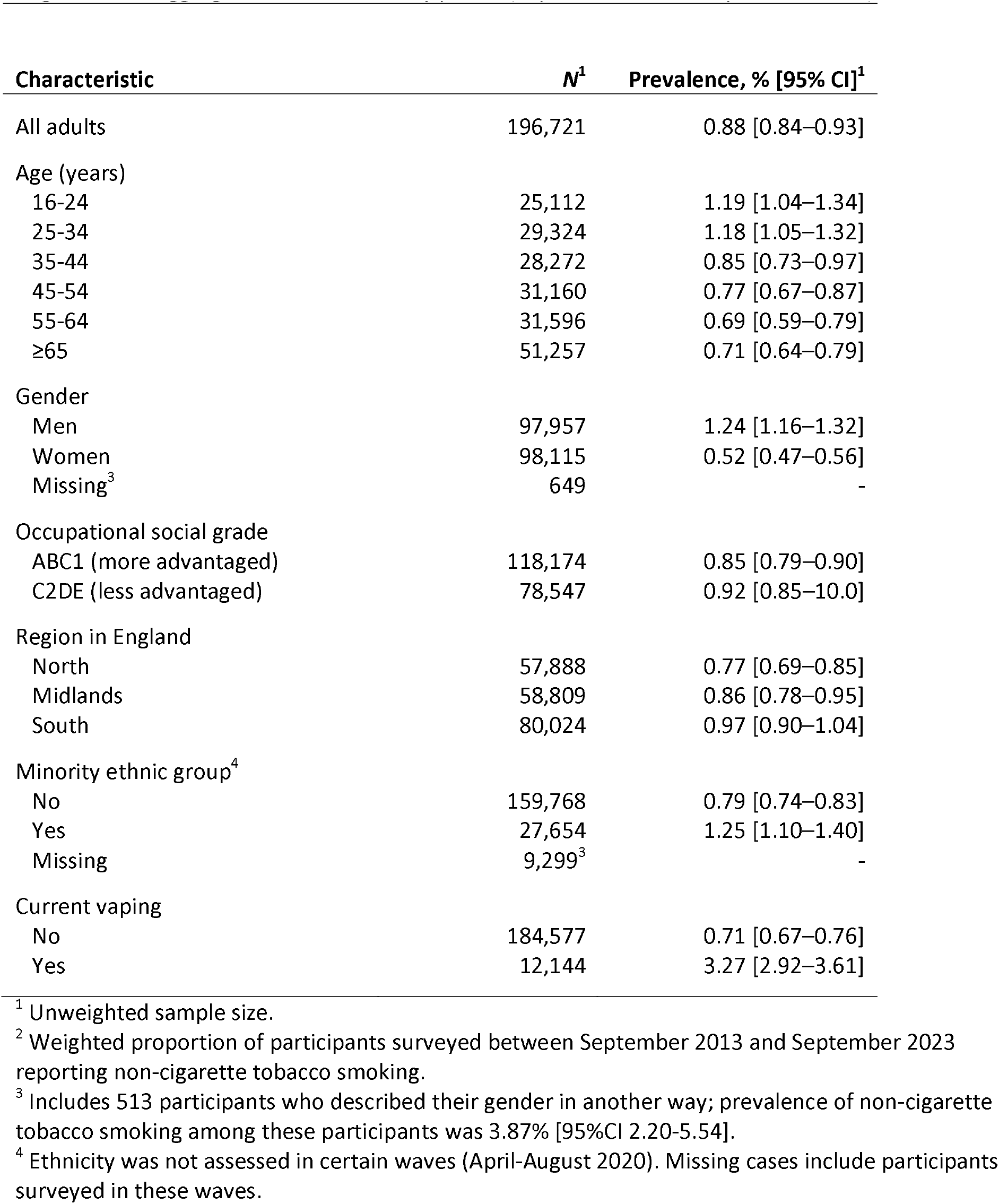
Unadjusted weighted prevalence of non-tobacco cigarette smoking among adults in England: data aggregated across the study period (September 2013 – September 2023)

| Characteristic | N<sup>1</sup> | Prevalence, % [95% CI]<sup>1</sup> |
| --- | --- | --- |
| All adults | 196,721 | 0.88 [0.84–0.93] |
| Age (years) |  |  |
| 16-24 | 25,112 | 1.19 [1.04–1.34] |
| 25-34 | 29,324 | 1.18 [1.05–1.32] |
| 35-44 | 28,272 | 0.85 [0.73–0.97] |
| 45-54 | 31,160 | 0.77 [0.67–0.87] |
| 55-64 | 31,596 | 0.69 [0.59–0.79] |
| ≥65 | 51,257 | 0.71 [0.64–0.79] |
| Gender |  |  |
| Men | 97,957 | 1.24 [1.16–1.32] |
| Women | 98,115 | 0.52 [0.47–0.56] |
| Missing<sup>3</sup> | 649 | - |
| Occupational social grade |  |  |
| ABC1 (more advantaged) | 118,174 | 0.85 [0.79–0.90] |
| C2DE (less advantaged) | 78,547 | 0.92 [0.85–10.0] |
| Region in England |  |  |
| North | 57,888 | 0.77 [0.69–0.85] |
| Midlands | 58,809 | 0.86 [0.78–0.95] |
| South | 80,024 | 0.97 [0.90–1.04] |
| Minority ethnic group<sup>4</sup> |  |  |
| No | 159,768 | 0.79 [0.74–0.83] |
| Yes | 27,654 | 1.25 [1.10–1.40] |
| Missing | 9,299<sup>3</sup> | - |
| Current vaping |  |  |
| No | 184,577 | 0.71 [0.67–0.76] |
| Yes | 12,144 | 3.27 [2.92–3.61] |

<sup>1</sup> Unweighted sample size.
<sup>2</sup> Weighted proportion of participants surveyed between September 2013 and September 2023 reporting non-cigarette tobacco smoking.
<sup>3</sup> Includes 513 participants who described their gender in another way; prevalence of non-cigarette tobacco smoking among these participants was 3.87% [95%CI 2.20-5.54].
<sup>4</sup> Ethnicity was not assessed in certain waves (April-August 2020). Missing cases include participants surveyed in these waves.

We reported the prevalence (with 95% confidence interval [CI]) of non-cigarette tobacco smoking, overall (aggregated across survey waves) and by survey year, among all adults and by age, gender, occupational social grade, region in England, ethnicity, and vaping status. For context, we also provided descriptive data on the proportion of smokers using cigarettes vs. exclusively using non-cigarette tobacco by survey year.

Trends in non-cigarette tobacco smoking prevalence over the study period were analysed using logistic regression with non-cigarette tobacco smoking as the outcome and time (survey month) modelled using restricted cubic splines with five knots (sufficient to accurately model trends across years without overfitting). This allowed for flexible and non-linear changes over time, while avoiding categorisation. We estimated the total number of adults in England smoking non-cigarette tobacco in September 2023 based on the most recent (2021) mid-year population estimates for England,^9^ and in September 2013 based on 2013 mid-year population estimates for England.^10^

To explore moderation of trends by age, gender, occupational social grade, region in England, ethnicity, and vaping status, we repeated the models including the interaction between the moderator of interest and time – thus allowing for time trends to differ across sub-groups. Each of the interactions was tested in a separate model. Age was modelled using restricted cubic splines with three knots (placed at the 5, 50, and 95% quantiles), to allow for a non-linear relationship between age and non-cigarette tobacco smoking.

We used predicted estimates from these models to plot the prevalence of non-cigarette tobacco smoking over the study period, among all adults and within each subgroup of interest. As age was modelled continuously, we displayed estimates for six specific ages (18-, 25-, 35-, 45-, 55-, and 65-year-olds) to illustrate how trends differ across ages. Note that the model used to derive these estimates included data from participants of all ages, not only those aged exactly 18, 25, 35, 45, 55, or 65 years. We also reported the absolute and relative changes across the whole time-series. Absolute changes were calculated as the percentage point change in prevalence (prevalence in September 2023 minus prevalence in September 2013) and relative changes were calculated as the prevalence ratio (PR; prevalence in September 2023 divided by prevalence in September 2013). We presented these alongside 95% CIs calculated using bootstrapping.

In an unplanned analysis, we used segmented regression to assess whether there was a step change in non-cigarette tobacco smoking at the start of the Covid-19 pandemic, which also coincided with the change from face-to-face to telephone interviews. We used logistic regression to model the trend in non-cigarette tobacco smoking before the pandemic (underlying secular trend; coded 1…*n*, where *n* was the total number of waves), the step-level change (coded 0 before the start of the pandemic in March 2020 and 1 after), and change in the trend (slope) post-onset of the pandemic relative to pre-pandemic (coded 0 before the pandemic and 1…*m* from April 2020 onwards, where *m* was the number of waves after the start of the pandemic). A linear pre-pandemic and pandemic trend was assumed, based on the relatively short length of the time-series (meaning we expected negligible differences between log-linear and linear trends). We used predicted estimates from these models to plot time trends in the prevalence of non-cigarette tobacco smoking alongside unmodelled quarterly data points.

## Results

A total of 197,299 (unweighted) adults aged ≥18 years were surveyed between September 2013 and September 2023. We excluded 578 (0.3%) with missing data on smoking status, leaving a final sample for analysis of 196,721 participants (weighted mean [SD] age = 47.9 [18.6] years; 50.8% female).

### Overall estimates of prevalence

Across the study period, the overall prevalence of non-cigarette tobacco smoking was 0.88% (**Table 1**). Groups with notably higher overall prevalence included younger adults, men and non-binary people, minority ethnic groups, and current vapers (**Table 1**). However, these overall estimates masked different patterns over time between subgroups of the population.

### Time trends

From September 2013 to September 2023, our primary (pre-registered) model indicated that the prevalence of non-cigarette tobacco smoking among all adults increased from 0.36% to 1.68% (PR=4.72 [95%CI 3.43–6.48]; **Table 2**). This equates to approximately 772,800 adult non-cigarette tobacco smokers in England in September 2023 (46 million adults^9^ x 1.68%); up from approximately 151,200 in September 2013 (42 million adults^10^ x 0.36%). As a result, the proportion of adult smokers in England who exclusively smoke non-cigarette tobacco is currently significantly higher than it was a decade ago (10.8% [9.64-12.0%] in 2022/23 vs. 2.25% [1.76-2.73%] in 2013/14; **Figure S1**).

**Table 2.** Modelled estimates of the change in prevalence of non-cigarette tobacco smoking among adults in England from September 2013 to September 2023.

| Characteristic | Prevalence, % [95%CI]<sup>1</sup> |  | Change in prevalence |  |
| --- | --- | --- | --- | --- |
|  | September 2013 | September 2023 | Relative change, prevalence ratio [95% CI]<sup>2</sup> | Absolute change, percentage points [95% CI]<sup>3</sup> |
| All adults | 0.36 [0.27–0.48] | 1.68 [1.45–1.94] | 4.72 [3.43–6.48] | 1.32 [1.07–1.60] |
| Age (years)<sup>4</sup> |  |  |  |  |
| 18 | 0.19 [0.08–0.45] | 3.21 [2.29–4.48] | 16.7 [7.62–47.9] | 3.02 [2.07–4.18] |
| 25 | 0.24 [0.13–0.43] | 2.61 [2.07–3.29] | 11.1 [6.39–23.3] | 2.38 [1.82–3.01] |
| 35 | 0.31 [0.21–0.46] | 1.97 [1.64–2.35] | 6.37 [4.28–10.3] | 1.66 [1.30–2.04] |
| 45 | 0.38 [0.25–0.59] | 1.53 [1.23–1.90] | 3.98 [2.57–6.66] | 1.14 [0.80–1.54] |
| 55 | 0.44 [0.28–0.67] | 1.26 [1.01–1.56] | 2.87 [1.85–4.94] | 0.82 [0.51–1.17] |
| 65 | 0.46 [0.32–0.65] | 1.09 [0.87–1.38] | 2.39 [1.60–3.76] | 0.64 [0.35–0.95] |
| Gender |  |  |  |  |
| Men | 0.69 [0.51–0.94] | 2.17 [1.80–2.61] | 3.13 [2.26–4.52] | 1.48 [1.04–1.94] |
| Women | 0.06 [0.03–0.13] | 1.07 [0.83–1.38] | 18.4 [9.06–52.5] | 1.01 [0.77–1.30] |
| Occupational social grade |  |  |  |  |
| ABC1 (more advantaged) | 0.40 [0.27–0.60] | 1.61 [1.35–1.92] | 4.00 [2.75–6.36] | 1.21 [0.94–1.52] |
| C2DE (less advantaged) | 0.30 [0.19–0.46] | 1.77 [1.39–2.26] | 5.94 [3.74–9.85] | 1.47 [1.06–1.95] |
| Region in England |  |  |  |  |
| North | 0.35 [0.22–0.56] | 1.44 [1.11–1.88] | 4.08 [2.78–8.99] | 1.09 [0.78–1.71] |
| Midlands | 0.36 [0.27–0.48] | 1.64 [1.41–1.91] | 4.60 [2.09–6.48] | 1.28 [0.64–1.52] |
| South | 0.36 [0.23–0.56] | 1.86 [1.52–2.28] | 5.18 [3.55–9.78] | 1.50 [1.19–2.01] |
| Ethnicity |  |  |  |  |
| White | 0.34 [0.25–0.47] | 1.67 [1.42–1.97] | 4.88 [3.52–7.14] | 1.33 [1.05–1.63] |
| Minority ethnic group | 0.40 [0.18–0.91] | 2.20 [1.59–3.04] | 5.49 [2.64–15.1] | 1.80 [1.08–2.62] |
| Using e-cigarettes |  |  |  |  |
| No | 0.32 [0.23–0.43] | 1.25 [1.05–1.48] | 3.95 [2.84–5.79] | 0.93 [0.70–1.17] |
| Yes | 1.25 [0.62–2.51] | 4.71 [3.60–6.14] | 3.76 [1.99–9.21] | 3.46 [2.01–4.89] |

CI, confidence interval.
<sup>1</sup> Data for September 2013 and September 2023 are weighted estimates of prevalence in these months (the first and last in the study period) from logistic regression with survey month modelled non-linearly using restricted cubic splines (five knots).
<sup>2</sup> Change in prevalence across the whole time-series (calculated as prevalence in September 2023 divided by prevalence in September 2013) with 95% confidence intervals calculated using bootstrapping.
<sup>3</sup> Absolute percentage point change in prevalence across the whole time-series (calculated as prevalence in September 2023 minus prevalence in September 2013) with 95% confidence intervals calculated using bootstrapping.
<sup>4</sup> Note that the model used to derive these estimates included data from participants of all ages, not only those who were aged exactly 18, 25, 35, 45, 55, or 65 years.
*Unmodelled estimates of prevalence within each survey year are provided in Table S1.*

The increase over time was not linear (**Figure 1**). Our primary model suggested prevalence was relatively stable up to February 2020, at an average of 0.46% [95%CI 0.40–0.53%], then increased sharply at the start of the Covid-19 pandemic, to 0.90% [0.82–0.99%] in March 2020. This was followed by a steadier rise, peaking at 1.97% [1.84–2.12%] in May 2022, before falling slightly to 1.68% [1.45–1.94%] by September 2023 (**Figure 1A**).

**Figure 1.**
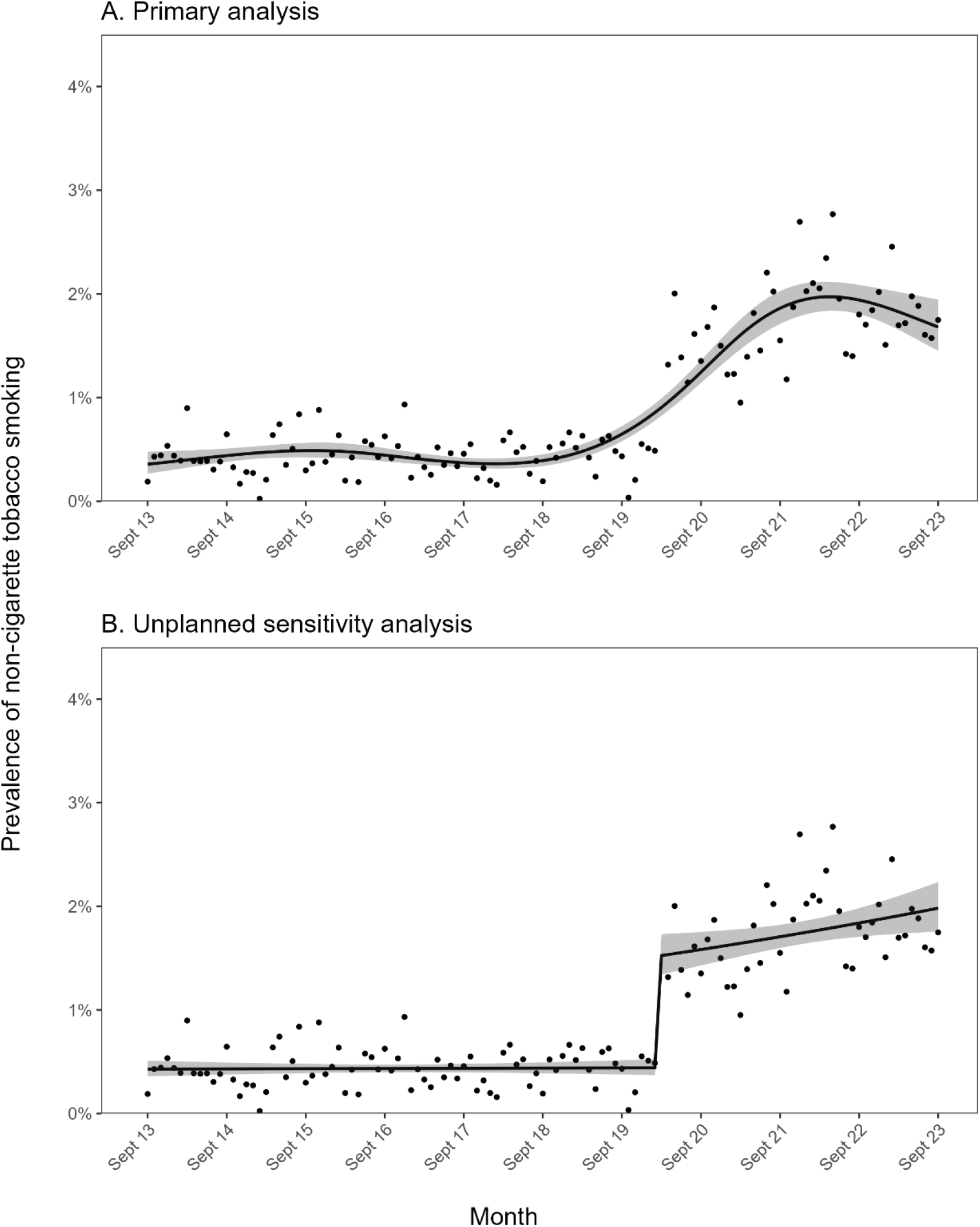
Trends in non-cigarette tobacco smoking among adults in England, September 2013 to September 2023. Panel A shows the results of the primary model, which modelled survey wave non-linearly using restricted cubic splines (five knots). Panel B shows the results of an unplanned sensitivity analysis, which used a segmented regression approach to model associations of the start of the Covid-19 pandemic with a step-level change in prevalence and a change in trend. Lines represent modelled weighted prevalence by monthly survey wave. Shaded bands represent 95% confidence intervals. Points represent unmodelled weighted prevalence by month.

We undertook an unplanned segmented regression analysis to explore the extent to which this pattern may have been driven by a step change at the start of the Covid-19 pandemic (March 2020), which also coincided with a change from face-to-face to telephone interviews, as opposed to the more gradual change in prevalence observed in our primary model. We observed a significant step-level increase in the prevalence of non-cigarette tobacco smoking at the start of the pandemic (OR_step-change_=3.475, 95%CI 2.783–4.338), from 0.44% [95%CI 0.37–0.52%] in February 2020 to 1.52% [1.34–1.73%] in March 2020 (**Figure 1B)**, which may have at least partially reflected the change in data collection. However, there was also a notable change in trend (OR_Δtrend_=1.006, 95%CI 1.000–1.013; **Figure 1B**). Before the pandemic, when data were collected face-to-face, prevalence of non-cigarette tobacco smoking was stable (monthly trend: OR_trend_=1.000, 95%CI 0.996–1.004). After the onset of the pandemic, when data were collected via telephone, prevalence of non-tobacco smoking increased by 0.6% per month (OR_trend_ x OR_Δtrend_ = 1.000 x 1.006 = 1.006) – or 7.2% per year (0.6% x 12 months; note these percentages represent the relative rather than absolute percentage point increase). This saw prevalence rise from 1.52% [1.34–1.73%] in March 2020 to 1.98% [1.76–2.23%] in September 2023.

This rise in prevalence of non-cigarette tobacco smoking was observed across all population subgroups to varying degrees (**Figure 2**). In particular, there were substantial differences by age (**Figure 2A**). In September 2013, prevalence of non-cigarette tobacco smoking was slightly lower at younger ages (e.g., 0.19% among 18-year-olds compared with 0.46% among 65-year-olds; **Table 2**). However, this pattern reversed over the subsequent decade as prevalence grew more rapidly and to higher levels among younger than older adults. As a result, in September 2023 prevalence of non-cigarette tobacco smoking was significantly higher at younger ages (e.g., 3.21% among 18-year-olds compared with 1.09% among 65-year-olds; **Table 2**).

**Figure 2.**
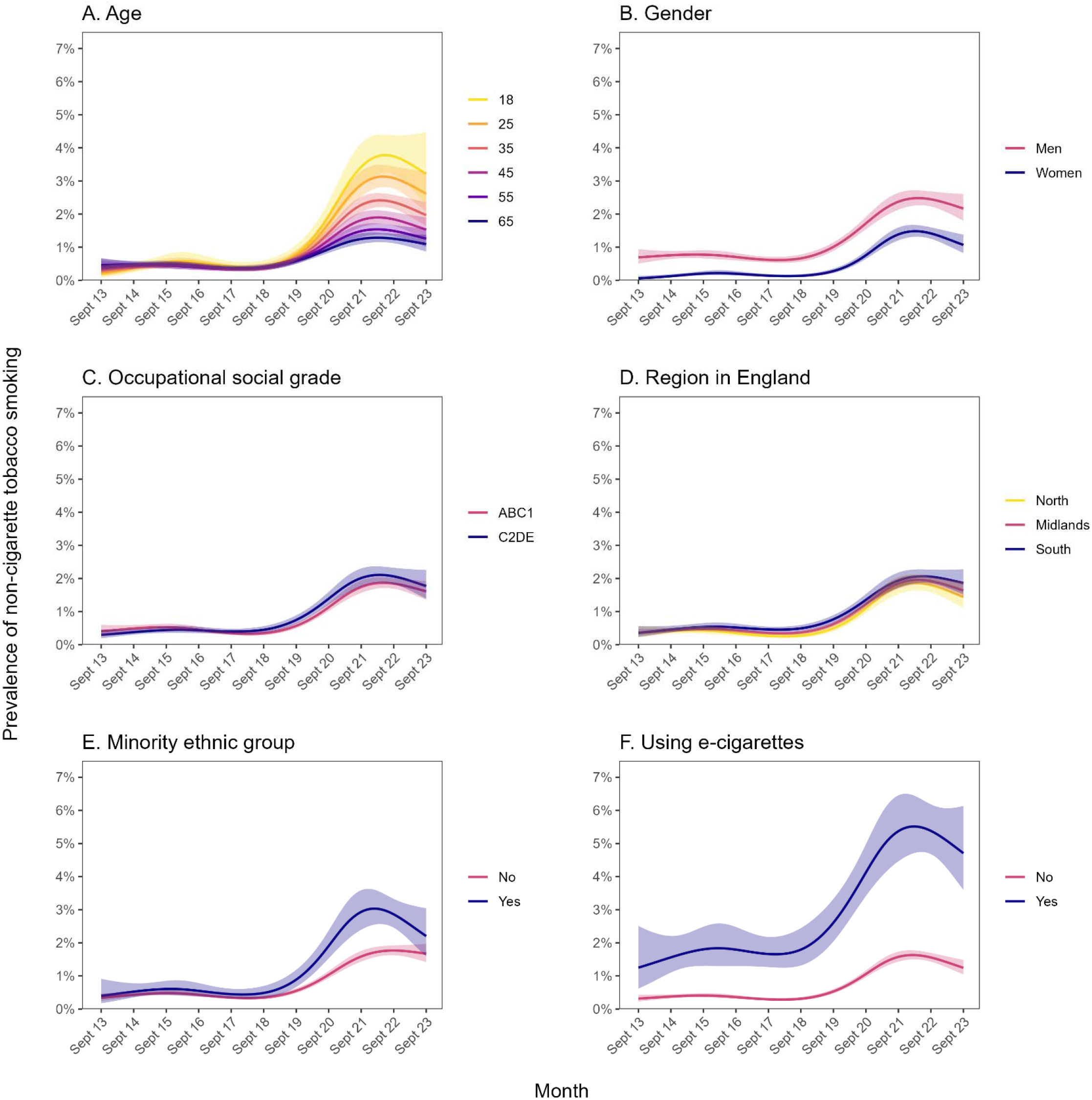
Trends in non-cigarette tobacco smoking within subgroups of adults in England, September 2013 to September 2023. Panels show trends by (A) age, (B) gender, (C) occupational social grade, (D) region in England, (E) ethnicity, and (F) vaping status. Lines represent modelled weighted prevalence by monthly survey wave, modelled non-linearly using restricted cubic splines (five knots). Shaded bands represent 95% confidence intervals.

Across the period, prevalence of non-cigarette tobacco smoking was significantly higher among men than women (**Table S1**), but time trends did not differ substantially (**Figure 2B**). The relative increase in prevalence was larger among women (PR=18.4 vs. 3.13 among men), who started from a very low baseline, but absolute changes over time were similar (**Table 2**).

Prevalence was similar across occupational social grades and regions in England, with no notable differences in changes over time (**Figure 2C-D**; **Table 2**).

Prevalence of non-cigarette tobacco smoking was similar across ethnic groups up to 2019, after which point patterns diverged (**Figure 2E**). The rise from 2020 onwards was larger among minority ethnic groups, peaking at 3.03% [95%CI 2.55–3.60] in February 2022 compared with a peak of 1.77% [95%CI 1.63–1.92] in September 2022 among white participants. The gap in prevalence narrowed between February 2022 and September 2023, with prevalence falling to 2.20% [95%CI 1.59–3.04] in September 2023 among minority ethnic groups but remaining relatively stable among white participants (1.67% [95%CI 1.42–1.97] in September 2023). As a result, changes from the start to the end of the time series were similar across ethnic groups (**Table 2**).

Across the period, non-cigarette tobacco smoking was significantly higher among vapers than non-vapers (**Table S1**). While the absolute change in prevalence was larger among vapers (+3.46 vs. +0.93 percentage points), the relative increase was similar by vaping status (**Table 2**).

## Discussion

Between September 2013 and September 2023, there was a non-linear increase in the prevalence of exclusive non-cigarette tobacco smoking among adults in England. While this rise in non-cigarette tobacco smoking was observed across all population subgroups, there was a significant age gradient whereby younger age was associated with a larger increase in prevalence. Across the decade, prevalence was consistently higher among men than women (as has been documented in other surveys^3,11^) and among current vapers than non-vapers, but was similar across occupational social grades and regions.

The timing of the rapid rise in prevalence of non-cigarette tobacco smoking we observed coincided with the onset of the Covid-19 pandemic in March 2020. It is possible this was driven by concerns about the health risks of smoking cigarettes. Early in the pandemic, there were concerns and studies suggesting that smoking might be associated with a higher risk of severe Covid-19 outcomes.^12–14^ Some people may have chosen to switch to non-cigarette combustible tobacco products, thinking that they could be less harmful or reduce their health risks.^15^ Consistent with this theory, there was a surge in quitting activity among smokers in England at the start of the pandemic,^6,7^ and cross-sectional data from the International Tobacco Control (ITC) Four Country Survey collected between February and June 2020 showed that a substantial proportion (11.8%) of recent ex-cigarette smokers in England, the United States, Canada, and Australia were using other combustible tobacco at this time.^11^

Another possible driver may have been economic factors. Many people have faced financial challenges due to the economic impact of the pandemic.^16^ Non-cigarette combustible tobacco products may have been perceived as more affordable options compared with traditional cigarettes, leading some people to switch to these alternatives. Alternatively, it is possible that a shift from in-person to online tobacco purchasing made consumers aware of the wider range of available products, or that home working was better suited to consumption of non-cigarette products or avoided concerns around social norms towards using them. It is also possible that some participants reporting non-cigarette tobacco use were referring to cannabis; there is evidence cannabis use may have increased during the pandemic.^17^

Concern for health as a motive for (switching to) non-cigarette tobacco smoking might be supported by the differences in the pattern of results we observed across ethnic groups. In the early stages of the pandemic, there was a particularly pronounced rise in non-cigarette tobacco smoking among minority ethnic groups, many of whom were at increased risk of severe Covid-19 outcomes.^18^ After the vaccination programme was rolled out and these risks were reduced, the prevalence of non-cigarette tobacco smoking among minority ethnic groups fell to levels comparable with white adults.

However, the rise in non-cigarette tobacco smoking since 2020 was also more pronounced among younger than older ages. This is the opposite pattern to what might be expected if the rise was being driven by concerns for health, given risks of adverse Covid-19 outcomes were greater at older ages.^19^ Cross-sectional data from the ITC Four Country Survey also documented higher levels of use of non-cigarette tobacco products among younger than older age groups in 2020.^11^ This pattern of results may reflect greater exploration of different products among younger adults.^11^ Over the same period when non-cigarette tobacco smoking increased, there was also a marked increase in vaping among adolescents and young adults,^20–22^ which may have prompted experimentation with other nicotine products. Across the study period, prevalence of non-cigarette tobacco smoking was higher among people who vaped. We did not capture frequency of product use, so our data do not provide any insight into patterns of use (i.e., regular daily use vs. experimental use) among those reporting non-cigarette tobacco smoking. Further research is needed to better understand the reasons non-cigarette tobacco smoking has increased since the start of the pandemic, the specific products being used, patterns of use, and how these differ across population subgroups.

Our findings have implications for policy. The UK Government plans to introduce a ‘smokefree generation’ policy, which would prohibit the sale of tobacco products in England to anyone born on or after 1 January 2009.^1^ Our data indicate that although cigarettes remain the product of choice for the vast majority of adult smokers in England, use of other combustible tobacco products (such as cigars, pipes, or shisha) has risen in recent years. As of September 2023, we estimate that ∼772,800 adults in England (more than one in every 10 smokers) do not use cigarettes at all but smoke other combustible tobacco products – five times more than the estimated number in September 2013 (∼151,200). The rise has been even more pronounced among younger ages. Collectively, these results suggest that while it is much less common than cigarette smoking, non-cigarette tobacco smoking is not negligible and possibly rising. The inclusion of these products in a smokefree generation policy is likely to be important for achieving the greatest reduction in youth uptake of tobacco smoking, as it would ensure young people who are unable to legally buy cigarettes do not buy other combustible tobacco products that are similarly harmful to health.^23,24^

Strengths of this study include the large, nationally representative sample and monthly data collection, which provides granular and up-to-date estimates of prevalence. There were also limitations. First, the survey did not capture the type of tobacco products smoked, so we are unable to determine whether the recent rise in non-cigarette tobacco smoking was driven by a particular product category. In addition, use of non-cigarette tobacco products was self-reported and there was potential for participants to misinterpret what is meant by ‘smoke tobacco of some kind’. While the question specifically stated that it was not referring to e-cigarettes, it is possible that the higher prevalence of non-cigarette tobacco use among current vapers partly reflected people misinterpreting the question to include e-cigarette use. Likewise, it is plausible that some participants may have considered cannabis use as another form of tobacco smoking. Further research is required to provide insight on the specific products driving the overall rise in non-cigarette tobacco use.

Another key limitation was the change in the mode of data collection (from face-to-face to telephone interviews) in April 2020 when the Covid-19 pandemic started, which coincided with the sharp rise in prevalence of non-cigarette tobacco smoking we observed. Previous studies have shown that estimates of substance use behaviour tend to be very similar across face-to-face and telephone interviews.^25–27^ Consistent with this, we collected data in parallel telephone and face-to-face surveys in March 2022 to evaluate the impact of this change on key sociodemographic and smoking parameters, and generally showed good comparability.^8^ However, the prevalence of non-cigarette smoking was 1.24 percentage points higher in the group surveyed via telephone than face-to-face (2.03% [95%CI 1.42-2.90] vs. 0.79% [0.48-1.31]),^8^ suggesting the change in methodology may account for some of the increase we observed. It is not clear how far this reflects a genuine effect of mode of data collection versus natural monthly variation in responses that would not be present if the data were collected over a longer period. Because non-tobacco smoking is relatively rare, there is a high degree of variability in the estimate for non-tobacco cigarette smoking between months even when the modality of assessment is constant (**Figure 1**). In any case, our data indicate the prevalence of non-cigarette tobacco smoking is currently substantially higher than previous estimates have suggested.

A third limitation is that we used a hybrid sampling approach rather than random probability sampling. However, comparisons with other sources suggest the survey recruits a nationally-representative sample and produces similar estimates of key smoking variables.^4,5^ Fourth, as a household survey, the sample excluded people living in institutions or experiencing homelessness, so our findings may not be representative of changes in non-cigarette tobacco smoking among these groups. Finally, our data do not offer any insight into use of non-combustible tobacco products (e.g., heated tobacco products), which are likely to have lower risks to health,^28^ but may also be included under the proposed smoke-free generation policy.

In conclusion, while the exclusive use of non-cigarette combustible tobacco remains rare among adults in England, it has increased in recent years. As of September 2023, there were approximately 772,800 adult non-cigarette tobacco smokers in England; around five times more than in September 2013. The prevalence of non-cigarette tobacco smoking has been consistently higher among men and current vapers. The rise in prevalence has differed by age, with a more pronounced rise leading to higher prevalence in September 2023 among younger than older ages.

## Data Availability

Data are available from the corresponding author on reasonable request.

## Declarations

### Ethics approval

Ethical approval for the STS was granted originally by the UCL Ethics Committee (ID 0498/001). The data are not collected by UCL and are anonymized when received by UCL.

### Competing interests

JB has received unrestricted research funding from Pfizer and J&J, who manufacture smoking cessation medications. LS has received honoraria for talks, unrestricted research grants and travel expenses to attend meetings and workshops from manufactures of smoking cessation medications (Pfizer; J&J), and has acted as paid reviewer for grant awarding bodies and as a paid consultant for health care companies. All authors declare no financial links with tobacco companies, e-cigarette manufacturers, or their representatives.

### Funding

Cancer Research UK (PRCRPG-Nov21\100002) funded the Smoking Toolkit Study data collection and SJ’s salary. For the purpose of Open Access, the author has applied a CC BY public copyright licence to any Author Accepted Manuscript version arising from this submission.

## Notes

### Clinical Protocols

https://osf.io/w7f3m/

