## Supplementary material for "Trends in non-cigarette tobacco smoking in England: a population survey 2013-2023": Figure S1

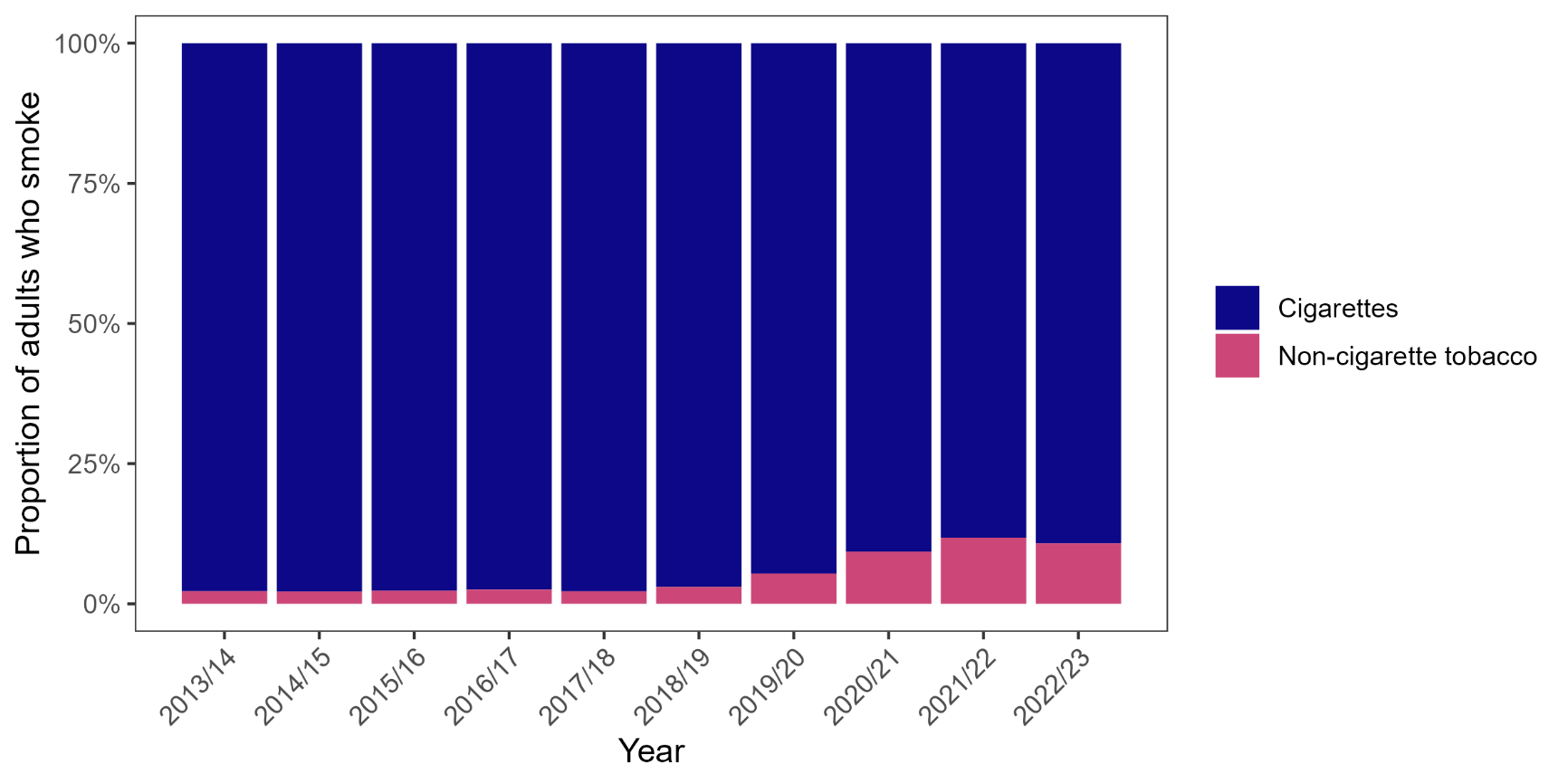

### **Figure S1. Proportion of adult smokers** in England who smoke non-cigarette tobacco, 2013/14 to 2022/23

The figure shows weighted data aggregated by year (September through August). Bars represent the proportion of smokers who smoke cigarettes vs. the proportion who exclusively smoke non-cigarette tobacco.

### Table S1. Prevalence of non-cigarette tobacco smoking by year among adults in England, 2013/14 to 2022/23

|  | **Prevalence, % [95% CI]^1^** | | | | | | | | | | |
| --- | --- | --- | --- | --- | --- | --- | --- | --- | --- | --- | --- |
|  | **Overall** | **2013/14** | **2014/15** | **2015/16** | **2016/17** | **2017/18** | **2018/19** | **2019/20** | **2020/21** | **2021/22** | **2022/23** |
| All adults | 0.88  [0.84–0.93] | 0.43  [0.34–0.53] | 0.42  [0.32–0.52] | 0.45  [0.35–0.55] | 0.45  [0.35–0.55] | 0.40  [0.31–0.49] | 0.49  [0.39–0.59] | 0.87  [0.72–1.03] | 1.56  [1.37–1.75] | 1.95  [1.73–2.16] | 1.82  [1.61–2.02] |
| Age (years) |  |  |  |  |  |  |  |  |  |  |  |
| 18-24 | 1.19  [1.04–1.34] | 0.36  [0.14–0.57] | 0.39  [0.13–0.65] | 0.72  [0.38–1.05] | 0.47  [0.19–0.74] | 0.46  [0.20–0.72] | 0.61  [0.27–0.95] | 0.78  [0.38–1.18] | 2.47  [1.75–3.19] | 3.15  [2.35–3.95] | 2.76  [2.0–3.51] |
| 25-34 | 1.18  [1.05–1.32] | 0.30  [0.11–0.5] | 0.29  [0.11–0.48] | 0.32  [0.13–0.52] | 0.53  [0.25–0.81] | 0.27  [0.07–0.47] | 0.56  [0.28–0.85] | 1.37  [0.85–1.88] | 2.28  [1.67–2.90] | 2.98  [2.30–3.66] | 2.87  [2.26–3.48] |
| 35-44 | 0.85  [0.73–0.97] | 0.50  [0.22–0.78] | 0.39  [0.10–0.68] | 0.32  [0.08–0.57] | 0.37  [0.13–0.61] | 0.31  [0.10–0.52] | 0.29  [0.09–0.50] | 1.22  [0.71–1.73] | 1.49  [0.99–1.99] | 2.15  [1.57–2.73] | 1.75  [1.22–2.27] |
| 45-54 | 0.77  [0.67–0.87] | 0.52  [0.26–0.79] | 0.54  [0.26–0.82] | 0.35  [0.13–0.58] | 0.40  [0.17–0.64] | 0.35  [0.14–0.55] | 0.29  [0.08–0.51] | 0.84  [0.49–1.19] | 1.33  [0.91–1.75] | 1.55  [1.10–2.00] | 1.67  [1.21–2.13] |
| 55-64 | 0.69  [0.59–0.79] | 0.35  [0.15–0.55] | 0.39  [0.15–0.63] | 0.48  [0.19–0.77] | 0.52  [0.26–0.77] | 0.39  [0.15–0.64] | 0.59  [0.30–0.88] | 0.50  [0.25–0.74] | 0.81  [0.48–1.15] | 1.29  [0.84–1.74] | 1.47  [1.02–1.91] |
| ≥65 | 0.71  [0.64–0.79] | 0.51  [0.31–0.70] | 0.47  [0.27–0.67] | 0.54  [0.35–0.73] | 0.45  [0.26–0.63] | 0.58  [0.38–0.78] | 0.59  [0.38–0.80] | 0.57  [0.35–0.80] | 1.30  [0.98–1.61] | 1.13  [0.81–1.44] | 0.91  [0.62–1.21] |
| Gender |  |  |  |  |  |  |  |  |  |  |  |
| Men | 1.24  [1.16–1.32] | 0.78  [0.59–0.96] | 0.70  [0.52–0.89] | 0.73  [0.55–0.90] | 0.67  [0.51–0.84] | 0.70  [0.53–0.86] | 0.84  [0.64–1.03] | 1.19  [0.93–1.45] | 2.16  [1.83–2.49] | 2.38  [2.03–2.73] | 2.28  [1.95–2.61] |
| Women | 0.52  [0.47–0.56] | 0.10  [0.04–0.16] | 0.14  [0.06–0.22] | 0.18  [0.09–0.27] | 0.23  [0.12–0.34] | 0.12  [0.05–0.19] | 0.15  [0.06–0.23] | 0.57  [0.39–0.75] | 0.96  [0.75–1.17] | 1.50  [1.24–1.76] | 1.29  [1.05–1.54] |
| Occupational social grade |  |  |  |  |  |  |  |  |  |  |  |
| ABC1 (more advantaged) | 0.85  [0.79–0.90] | 0.49  [0.35–0.63] | 0.39  [0.27–0.52] | 0.53  [0.38–0.67] | 0.45  [0.32–0.58] | 0.38  [0.27–0.49] | 0.47  [0.35–0.60] | 0.64  [0.48–0.79] | 1.51  [1.28–1.74] | 1.79  [1.55–2.04] | 1.78  [1.54–2.02] |
| C2DE (less advantaged) | 0.92  [0.85–10.0] | 0.36  [0.24–0.48] | 0.45  [0.29–0.60] | 0.35  [0.22–0.48] | 0.46  [0.31–0.61] | 0.43  [0.28–0.57] | 0.51  [0.34–0.69] | 1.17  [0.88–1.46] | 1.62  [1.30–1.95] | 2.14  [1.76–2.53] | 1.86  [1.51–2.21] |
| Region in England |  |  |  |  |  |  |  |  |  |  |  |
| North | 0.77  [0.69–0.85] | 0.42  [0.26–0.57] | 0.31  [0.17–0.45] | 0.49  [0.30–0.69] | 0.36  [0.19–0.53] | 0.33  [0.18–0.47] | 0.31  [0.17–0.46] | 0.73  [0.48–0.98] | 1.30  [0.98–1.62] | 1.87  [1.46–2.27] | 1.72  [1.35–2.09] |
| Midlands | 0.86  [0.78–0.95] | 0.40  [0.24–0.57] | 0.53  [0.32–0.75] | 0.24  [0.10–0.37] | 0.36  [0.20–0.51] | 0.43  [0.25–0.61] | 0.50  [0.31–0.69] | 0.83  [0.57–1.10] | 1.68  [1.31–2.05] | 2.10  [1.68–2.52] | 1.58  [1.24–1.92] |
| South | 0.97  [0.90–1.04] | 0.46  [0.30–0.62] | 0.41  [0.26–0.56] | 0.57  [0.40–0.74] | 0.59  [0.41–0.76] | 0.43  [0.29–0.57] | 0.60  [0.42–0.78] | 0.99  [0.72–1.26] | 1.65  [1.34–1.96] | 1.88  [1.57–2.20] | 2.05  [1.71–2.38] |
| Minority ethnic group |  |  |  |  |  |  |  |  |  |  |  |
| No | 0.79  [0.74–0.83] | 0.42  [0.32–0.52] | 0.40  [0.30–0.51] | 0.43  [0.32–0.53] | 0.45  [0.34–0.55] | 0.38  [0.29–0.47] | 0.47  [0.36–0.58] | 0.34  [0.20–0.48] | 1.35  [1.16–1.53] | 1.77  [1.55–2.00] | 1.71  [1.49–1.93] |
| Yes | 1.25  [1.10–1.40] | 0.52  [0.22–0.83] | 0.53  [0.24–0.83] | 0.58  [0.28–0.88] | 0.45  [0.20–0.71] | 0.51  [0.22–0.81] | 0.61  [0.29–0.93] | 0.59  [0.18–0.99] | 2.70  [1.93–3.46] | 2.98  [2.29–3.67] | 2.45  [1.89–3.00] |
| Current vaping |  |  |  |  |  |  |  |  |  |  |  |
| No | 0.71  [0.67–0.76] | 0.38  [0.29–0.47] | 0.36  [0.26–0.45] | 0.36  [0.27–0.46] | 0.36  [0.27–0.45] | 0.34  [0.26–0.43] | 0.36  [0.27–0.46] | 0.73  [0.58–0.88] | 1.36  [1.18–1.54] | 1.59  [1.39–1.79] | 1.41  [1.22–1.61] |
| Yes | 3.27  [2.92–3.61] | 1.43  [0.68–2.17] | 1.55  [0.67–2.43] | 1.76  [0.96–2.56] | 2.02  [1.04–2.99] | 1.37  [0.65–2.09] | 2.71  [1.65–3.77] | 3.22  [2.00–4.45] | 4.20  [2.91–5.50] | 5.87  [4.55–7.19] | 4.98  [3.98–5.98] |

^1^ Data are weighted proportions, overall (across all participants surveyed between September 2013 and September 2023) and aggregated by survey year (September through August).
